# Impact of Delta and Vaccination on SARS-CoV-2 transmission risk: Lessons for Emerging Breakthrough infections

**DOI:** 10.1101/2022.03.02.22271385

**Authors:** Kalpana Sriraman, Ambreen Shaikh, Smriti Vaswani, Tejal Mestry, Grishma Patel, Shalini Sakthivel, Vikas Oswal, Pratibha Kadam, Kayzad Nilgiriwala, Daksha Shah, Mangala Gomare, Nerges Mistry

## Abstract

With the continuous emergence of SARS-CoV-2 variants of concern and implementation of mass-scale interventions like vaccination, understanding factors affecting disease transmission has critical implications for control efforts. Here we used a simple adapted N95 mask sampling method to demonstrate the impact of circulating SARS-CoV-2 variants and vaccination on 92 COVID-19 patients to expel virus into the air translating to a transmission risk. Between July and September 2021, when the Delta was the dominant circulating strain in Mumbai, we noted a two-fold increase in the proportion of people expelling virus (95%), about an eighty-fold increase in median viral load and a three-fold increase in high emitter type (41%; people expelling >1000 viral copy numbers in 30 minutes) compared to initial strains of 2020. Eight percent of these patients continued to be high emitters even after eight days of symptom onset, suggesting a probable increased transmission risk for Delta strain even at this stage. There was no significant difference in expelling pattern between partial, full and un-vaccinated individuals suggesting similar transmission risk. We noted significantly more infections among vaccinated study patients and their household members than unvaccinated, probably due to increased duration from vaccination and/or increased risk behaviour upon vaccination due to lower perceived threat. This study provides biological evidence for possible continued transmission of the Delta strain even with vaccination, emphasizing the need to continue COVID-19 appropriate behaviour. The study also indicates that the mask method may be useful for screening future vaccine candidates, therapeutics or interventions for their ability to block transmission.

## Introduction

The year 2021 was marked by the spread of the SARS-CoV-2 Delta variant and unprecedented efforts to vaccinate the global population. India approved two vaccines, Covishield (Adenovirus vector vaccine, ChAdOx1 nCoV-19-Serum Institute of India) and COVAXIN (Inactivated whole virus vaccine, BBV152-Bharat Biotech), in January 2021. The emergence of SARS-CoV-2 variants like the Delta raised concerns regarding vaccine efficacy and its effectiveness against transmission. As infections increased, it became clear that the Delta strain dominated the vaccine breakthrough cases (1, 2). While the overall efficacy of the approved Indian vaccines was low for Delta infections, their effectiveness in preventing severe infections ranged from 70% - >90% depending on the population (3-5).

From a pandemic control perspective, although understanding vaccine effectiveness is important in terms of risk of infection and severe disease, it is also critical to understand the impact of vaccination on preventing transmission. However, clinical trials conducted to measure vaccine effectiveness are usually not designed to measure the effect of the vaccine on disease transmission. Such information gets generated only after introducing vaccines into the population through real-world observational studies. Thus, data on vaccine efficacy on transmission are limited. Few studies have reported the measurement of vaccine efficacy against Delta transmission through household contact tracing. These were predominantly reported from western countries and/or on the effectiveness of the mRNA vaccines (6-8). Such data are limited for other vaccines types, except for one UK study, which demonstrated a marginal but statistically non-significant benefit of the AZD1222 (ChAdOx nCoV19 vaccine) on secondary infections and transmission from Delta index cases (9). More frequently, indirect measures like viral load in an individual’s nasopharyngeal swab (NPS) samples are used as surrogate markers to gauge the vaccine’s effects on the transmission (10). It is hypothesized that the vaccines are likely to reduce transmission risk if they can reduce an individual’s viral load. However, emerging evidence shows marginal differences in viral load between vaccinated and unvaccinated individuals, especially with the Delta infections, although viral load decline with time appears to be faster in vaccinated individuals (7, 9, 11).

Since the predominant transmission route for SARS-CoV-2 is through respiratory aerosols and droplets, it is reasonable to assume that an infected person must expel the virus in respiratory particles for transmission to occur. Our work in COVID-19 patients showed that measuring viral load in respiratory particles expelled by patients while talking, coughing, and breathing collected using a non-invasive adapted N-95 mask sampling could be a more useful indicator for assessing individual transmission risk (12). We found that the viral load in the expelled respiratory particles represented known transmission rates in the community better than the Ct values estimated in NPS samples. This distinction prompted us to use the mask sampling method to understand the potential individual transmission risk of circulating strains in vaccinated individuals at early and late stages of the SARS-C0V-2 infection.

Here we report the results of our prospective observational study conducted in Mumbai in the wake of the Delta dominated the second wave of SARS-CoV-2 infection. The primary aim of this study was to understand the effect of circulating strains and vaccination (Covishield and COVAXIN) on the ability of COVID-19 patients to expel the virus in respiratory particles at early and late stages of infection, thereby determining their potential transmission risk. We further report the risk of infection among vaccinees and discuss the factors that may influence it.

## Methods

### Patient Recruitment and Sample Collection

The study was undertaken between July - September 2021 after the approval by the Institutional Research Ethics Committee at the Foundation for Medical Research (FMR/IREC/C19/01/2021; Clinical trials registry No CTRI/2021/07/035143). A total of 95 vaccinated and unvaccinated COVID-19 patients residing in Mumbai were recruited through referrals from study collaborators in the public and private systems. Eligible participants were consenting COVID-19 patients, 18 years of age or older, who had mild disease with SpO2 ≥95 at room air, not on supplemental oxygen at the time of recruitment, and fit for mask sampling. The patients were primarily recruited via phone call within 24 hours of diagnosis. Patient enrolment involving written consent and first sample collection was carried out within 48 hours of the RT-PCR diagnosis. Depending on their vaccination status, the patients were assigned to the following five groups: -

1. One dose Covishield vaccinated (**Cs-D1**, n=26)
2. Two doses Covishield vaccinated (**Cs-D2**, n=29)
3. One dose COVAXIN vaccinated (**Cx-D1**, n=2)
4. Two doses COVAXIN vaccinated (**Cx-D2**, n = 25)
5. Unvaccinated (n=13)

Mask and NPS samples were collected from all patients at two-time points - first sample collection within 48 hours of COVID-19 positive report, and the second sample collection (follow-up sample) between 8-12 days from the first reported COVID-19 symptom (or from the date of diagnosis for asymptomatic positives). Demographic characteristics, clinical presentation, treatment, and household information, including their vaccination and infection status, were recorded for all the study participants at all interaction points. A final telephonic follow-up was carried out between 15-21 days from the first reported symptom to document the patients’ health outcome and information on any subsequent household infections. The mask sample collection involved collecting expelled respiratory particles of patients using a modified N95 mask in a 30-minute process that included 20-minute undirected-10-minute directed vocal and breathing manoeuvres followed by calculation of sampling score based on the intensity of tasks performed as previously described (12). At the end of the sample collection process, the membrane attached to the inner surface of the modified mask was dissolved immediately in RNAzol™ (Sigma-Aldrich, MO, USA). Immediately after mask sampling, the swab sample was collected in viral transport media (VTM; Vi-Trans, Cellkraft Biotech Pvt. Ltd, Bengaluru, India). Both samples were transported to the FMR laboratory and stored for not more than 24 hours at 4^0^C before further processing.

### Sample Processing and RT-PCR

RNA was extracted from VTM using QiaAmp viral RNA mini kit (Qiagen GmBH, Hilden, Germany) as per the manufacturers’ protocol, while RNA from RNAzol™ was extracted as previously described, combining RNAzol extraction with QIAmp kit purification (12). The RT-PCR was carried out in CFX 96 real-time thermal cycler (Bio-Rad Laboratories, California, USA), with COVIPath™ COVID-19 Kit (Invitrogen Bio Services India Pvt Ltd., Bengaluru, India) as per the manufacturer’s protocol. The kit detects the N and O genes specific for SARS-CoV-2. COVID-19 negative NPS samples and RNA isolated from tuberculosis patient mask samples collected before December 2019 (Pre-COVID) were used as negative controls. As the samples were from RT-PCR confirmed COVID-19 patients, detection of both N and O genes or the O gene with visible sigmoidal PCR amplification curves and detectable Ct value (<40 as against <35 used for diagnosis) were considered positive. A standard curve was generated by performing 10-fold serial dilutions of the commercially available IVT RNA kit (Taqpath COVID-19 Control kit, Thermo Fischer Scientific, USA) to determine the viral load (RNA copy numbers) in SARS-CoV-2 positive samples. In mask samples, viral copies of >1000 in the 30-minute collection time defined the patient as a high emitter with a potential increased risk for transmission (12). A viral copy number of 100-999 defined them as moderate emitters with moderate transmission risk, while patients with <100 copy numbers were considered low emitters with minimal transmission risk. All samples with Ct value <33 (n=83) were subjected to whole-genome sequencing (WGS) by the Oxford Nanopore sequencing using MinION (13), and viral lineage was determined in 75/83 samples using the PANGOLIN tool (*v3*.*1*.*17)* (14).

### Household Infection Rate and Risk of Infection Calculations

For each patient, the number of household members, their infection status, date of diagnosis if infected and vaccination status were mapped. The household infection rate was calculated as follows: Household infection rate (HHR) = (number of confirmed positive members in the household at enrolment + number of new positive members at follow-up + number of new positive members at final telephonic follow-up)/total number of individuals living in the same household. For the risk of infection calculation, both patients’ and their household members’ data were combined and stratified based on their infection status and vaccination status. COVID-19 positivity rate was calculated for each vaccine status, and the percentage positivity was indicated as the risk of infection.

### Statistical analysis

Results were statistically analyzed using Graph Pad Prism software (version 9). Percentages were calculated for categorical variables, and Fisher exact test was applied. For continuous variables, the median with interquartile range (IQR) and Mann Whitney unpaired test were applied. A p-value of < 0.05 was considered significant for both tests. Multivariate logistic regression analysis was used to understand the interaction of various factors on the emission pattern of the SARS-CoV-2 virus and the household infection pattern. Where necessary, a power analysis was carried out to evaluate the impact of sample size on the reported results using the Openepi tool (15). Probit modelling was performed with MedCalc version 20.019 (MedCalc Software Ltd) for evaluating the probability of detecting the SARS-CoV-2 virus in mask and swab samples on a particular day in the infection period.

## Results

Of the 95 confirmed COVID-19 patients enrolled in the study, three patients were NPS and mask RT-PCR negative at the first sample collection and thus were excluded from the primary analysis. A total of 84 patients provided both enrolment and follow-up samples. Follow up samples could not be collected from a few patients due to patient refusal or admission to a hospital beyond the study jurisdiction. Overall, there were 55.5% male and 44.5% female. The median age of the recruited patients was 40 (IQR 30-50, Table 1). Of the 92 patients analyzed, 89 patients had a mild form of the disease as per the disease severity definition of the Govt. of India (16). For three patients, their disease status changed after enrolment into the study, and they were shifted to a hospital with final disease status classified as moderate. We grouped the 92 patients’ data into five groups based on the vaccination status mentioned in the Methods and compared their demographics, COVID-19 clinical characteristics, mask sampling scores (Table 1).

**Table 1.**
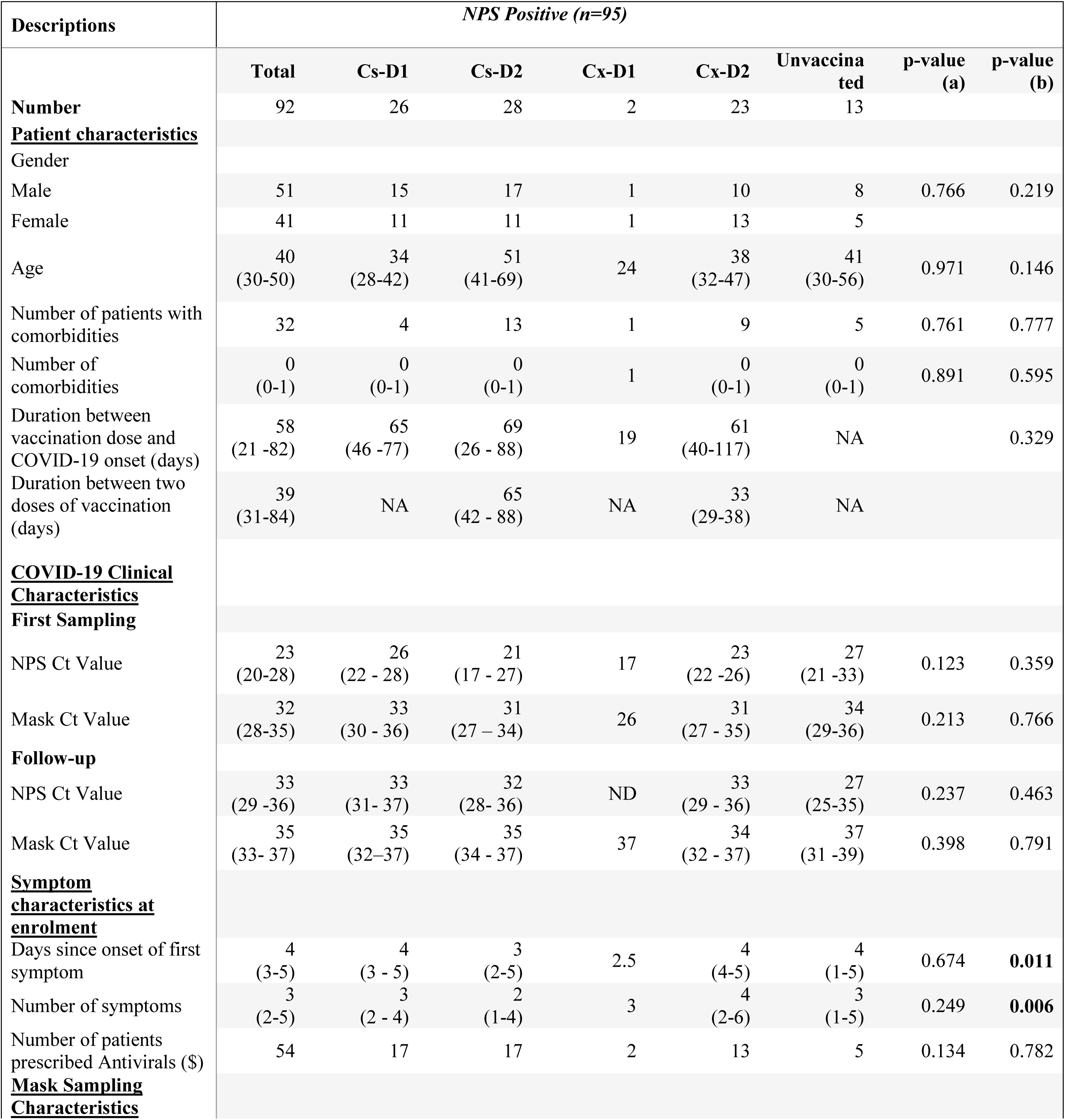
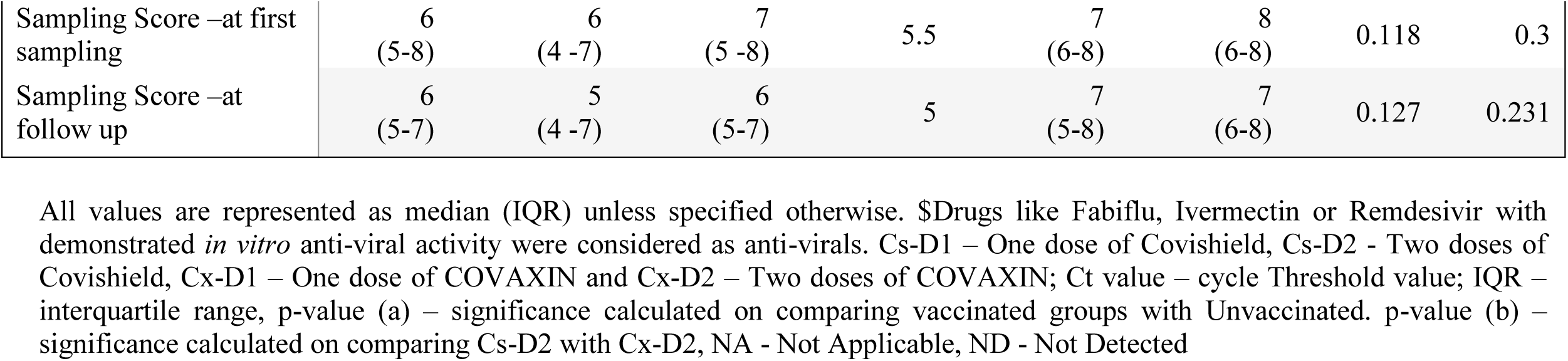
Comparison of patient demographics, COVID-19 clinical characteristics and mask sampling characteristics at enrolment and follow-up stratified based on vaccination status

Most demographic and baseline clinical characteristics were similar between the groups except for age and the number of COVID-19 symptoms reported at the time of recruitment (Table-1, Supplementary Table 1). The median age of patients in the Cs-D2 group (51) was higher than the other groups, probably due to the older age group achieving full vaccination status earlier than the younger age group as per the prevalent vaccination policy and Covishield availability in the initial phases of the vaccine rollout in the city. In addition, days since first symptom onset and a median number of symptoms reported at enrolment were significantly higher in the Cx-D2 group than in Cs-D2 (p-value < 0.05, Table 1). WGS data analysis confirmed that the SARS-CoV-2 positive samples were either Delta (n= 66, 88%) or Delta derivative (n=9, Supplementary Table 2).

### Mask positivity and Emission pattern in mask samples at enrolment

We compared the proportion of patients expelling the virus (mask positivity) based on vaccination status (Table 2). As the Cx-D1 group had only two patients, it was excluded from the comparison. All patients in the vaccinated groups (partial/full Cs/Cx) were NPS positive at the first sampling, and Ct values were similar between the vaccine groups. The overall proportion of patients expelling the virus was 95% and was comparable between partially, fully vaccinated, and unvaccinated groups (Table 2). All patients with NPS Ct value <30 was 100% mask positive, while the mask positivity reduced to 61% for NPS Ct value >30 (Supplementary Figure 1A). In the current analysis, the 95% mask positivity observed among COVID-19 patients was 2.3-fold more than the mask positivity rate we recorded during the first wave of COVID-19 in Mumbai (42% positivity, (12)).

**Table 2.** Comparison of mask and NPS positivity at enrolment and follow-up

| *Stage* | *This Study* |  |  |  |  |  |  | *Previous Study (2020)* |
| --- | --- | --- | --- | --- | --- | --- | --- | --- |
|  | Category | Cs-D1 | Cs-D2 | Cx-D1 | Cx-D2 | Unvaccinated | Overall | Unvaccinated 1st Wave |
|  | **Total Enrolled** | **26** | **29** | **2** | **25** | **13** | **95** |  |
| *Enrolment* | Total patients after exclusion | 26 | 28 | 2 | 23 | 13 | 92 | 31 |
|  | Swab Positives | 26 | 28 | 2 | 23 | 12 | 91 | 29 |
|  | Swab positivity (%) | **100** | **100** |  | **100** | **92** | **99** | **94** |
|  | Mask Positives | 25 | 26 | 2 | 22 | 12 | 87 | 13 |
|  | Mask Positivity (%) | **96** | **93** |  | **96** | **92** | **95** | **42** |
|  | High Emitters (%) | **27** | **50** | **50** | **48** | **33** | **41** | 13 |
| *Follow-up* | Lost to Follow-up | 1 | 2 | 1 | 3 | 1 | 8 |  |
|  | Total Followed-Up | 25 | 26 | 1 | 20 | 12 | 84 |  |
|  | Swab Positives | 21 | 24 | 0 | 17 | 10 | 72 |  |
|  | Swab Positivity (%) | **84** | **92** |  | **85** | **83** | **86** |  |
|  | Mask Positives | 13 | 16 | 1 | 18 | 7 | 55 |  |
|  | Mask Positivity (%) | **52** | **62** |  | **90** | **58** | **65** |  |
|  | High Emitters % | 8 | 4 | 0 | 15 | 8 | 8 |  |

Cs-D1 – One dose of Covishield, Cs-D2 - Two doses of Covishield, Cx-D1 – One dose of COVAXIN and Cx-D2 – Two doses of COVAXIN; Ct value – cycle Threshold value; IQR – interquartile range. The previous study – results are available at https://doi.org/10.1371/journal.pone.0249525. Percentages are indicated in bold

We then compared the Ct values and calculated the viral load in these samples. Overall, we observed a 5-Ct value decrease (≈80-fold increase in viral load) in the current cohort (Median 32 IQR 28-35 in mask and Median 23 IQR 21-28 in NPS) compared to the previous year 2020 infections (37 IQR 33-38-mask and 29 IQR 24-31-NPS). On stratifying patient data into high, moderate and low emitters as defined in the methods section, the overall percentage of high emitters in the current study was 3.3-fold more than the percentage observed in the previous study cohort (Figure 1A) (41%-present study v/s 13%-(12)), supporting the higher transmission of SARS-CoV-2 observed during the second wave in 2021. However, no significant difference was observed between the vaccine groups in the emission pattern (Figure 1B). Our analysis showed that high emitter patients were more likely to have high NPS viral load (p<0.0001), cough as a symptom (p=0.038), and shorter duration between symptom onset and enrolment (p=0.023, Supplementary Table 3). After adjusting for confounding factors, multivariate logistics regression analysis showed that only NPS viral load (p=0.027) influenced the high emitter pattern (Supplementary Table 3). However, linear regression analysis showed that the observed association of NPS viral load with mask viral load was weak (R^2^ = 0.1427 – high emiiters and 0.457 – mask positives).

**Figure 1.**
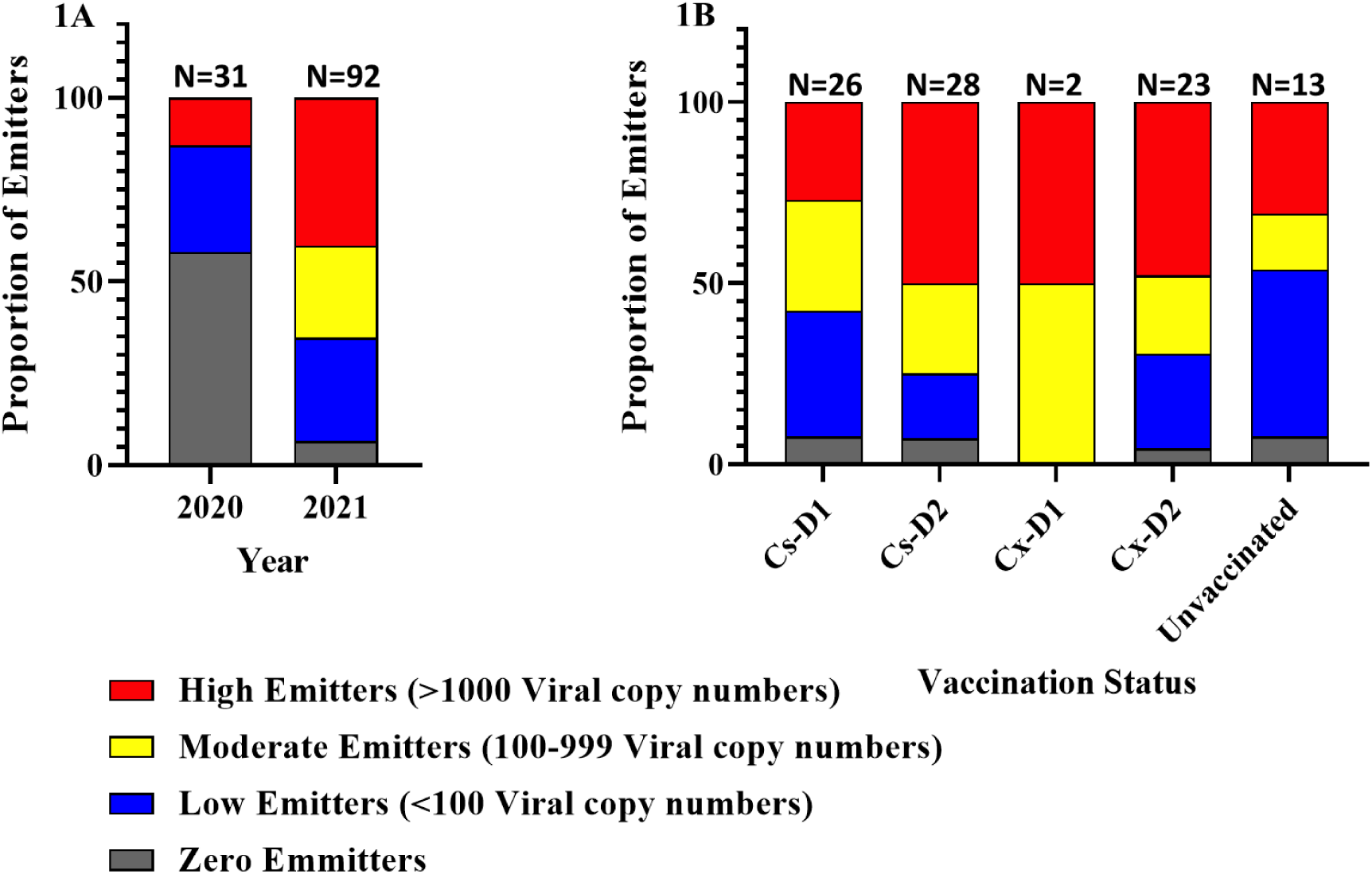
**1A** – Comparison of the proportion of patients classified as high emitters, moderate emitters, low emitters and zero emitters between the first wave (2020,(12)) and this study (2021). **1B** - Comparison of the proportion of patients classified as high emitters, moderate emitters, low emitters and zero emitters based on the vaccination status. Cs-D1-Covishield Dose 1, Cs-D2-Covishield Dose 2, Cx-D1-COVAXIN Dose 1 and Cx-D2-COVAXIN Dose 2

### Mask positivity and Emission pattern in mask samples at follow-up stage

We further observed that even after eight days of symptom onset, the mask positivity rate remained high, although less than swab positivity, for patients in all the groups (Table 2). Among the vaccinated groups, the patients in the Cx-D2 group had significantly higher mask positivity (p<0.05) at eight days (90%) compared to Cs-D1 or Cs-D2 group patients, indicating that COVAXIN vaccinated patients may be clearing the virus slower than the other groups. However, this significance had a power of only 58% for the sample size. The viral load data showed that the mask viral load did not correlate to NPS viral load (Supplementary Figure 1B), and only patients in Cs-D1and Cs-D2 groups had a significant reduction in viral load (p<0.01) at follow-up (Figure 2). Although most patients showed a decrease in viral load and shifted from high or moderate emitters to low emitters or zero emitters (mask negative), about 8% of the patients continued to be high emitters even after eight days of symptom onset. Statistical analysis showed the high emitters at follow-up were more likely to continue having cough as a symptom (OR=8.323, p-value =0.0434). In order to predict the expelling pattern and understand potential transmission risk on the tenth day of symptom onset - when general recommendations advised the ending of patient isolation-probit modelling of the data was carried out. The probability of being mask positive decreased from the predicted 100% on day 1 to 64.9% on the tenth day (Supplementary Figure 2A), while the swab positivity only reduced to 83% (Supplementary Figure 2B). More importantly, the probability of being a high emitter significantly reduced from predicted 90% on day 1 to 7% on day 10 (Supplementary Figure 2C)

**Figure 2.**
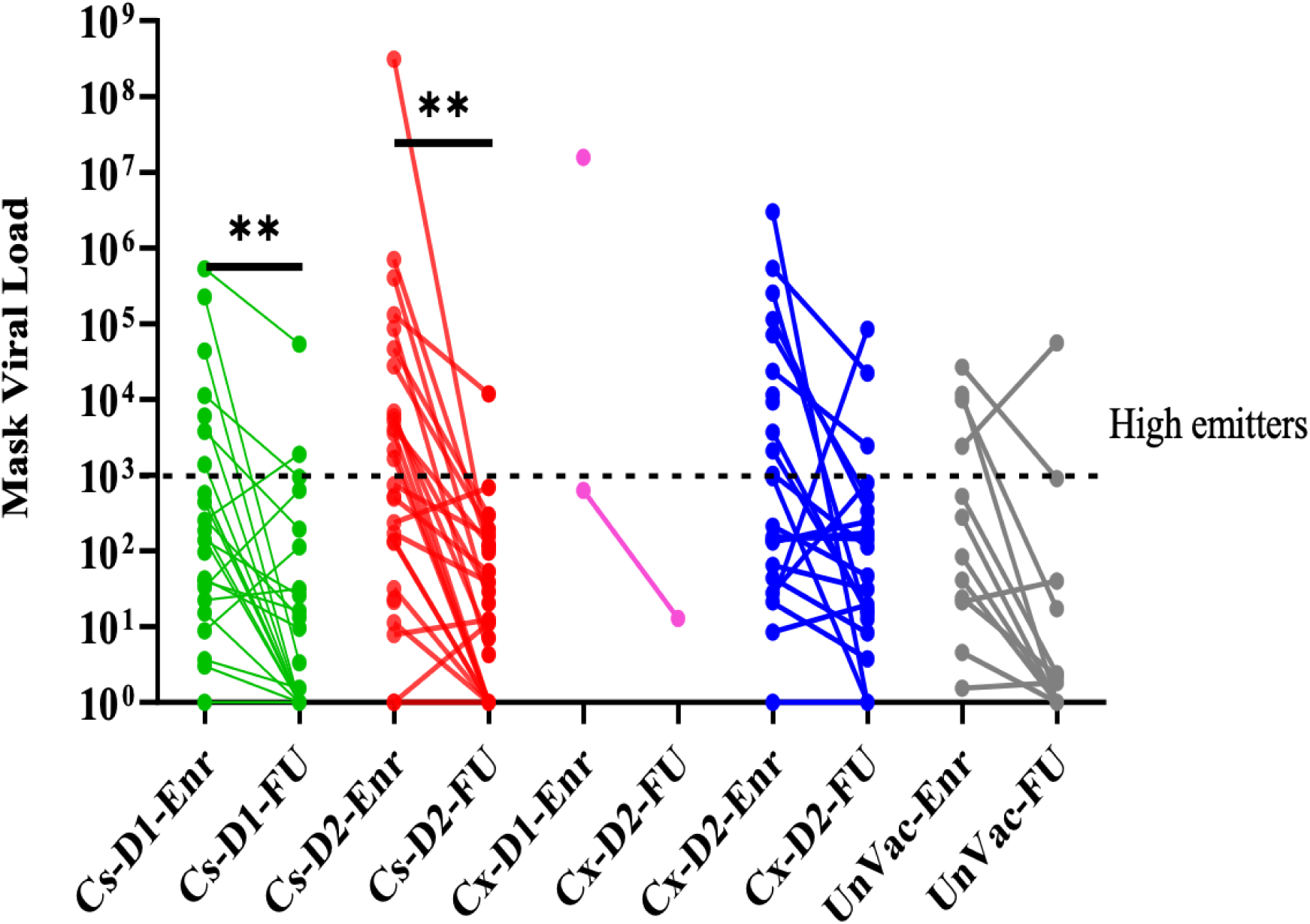
Mask viral load reduction from enrolment (Enr) to follow-up (FU) in the various vaccine groups, Cs-D1-Covishield Dose 1, Cs-D2-Covishield Dose 2, Cx-D1-COVAXIN Dose 1 and Cx-D2-COVAXIN Dose 2, UnVac-Unvaccinated, Enr-Enrolment and FU-Follow up. ** indicates statistical significance with p<0.01

### Vaccination and Risk of Infection in Households

To understand the possible link between vaccination, mask viral load and transmission, we estimated the household infection rate as described in the methods. Since many household members, including study patients, tested positive at the same time or within two days of one-member testing positive, we could neither define study patients as an index nor establish the causal relationship between their viral load on household infection rates. Nevertheless, the median HHR was significantly higher in Cs-D1 (Median 67%, p < 0.01) and Cs-D2 (Median 88%, p < 0.001) groups compared to the unvaccinated (Median 20%) or Cx-D2 group (Median 50%) (Figure 3). To further verify, we re-stratified the households as single infection (only study patient) and multiple infections (study patient + family member) households and determined the influencing factors by multivariate analysis. Multiple infections in households were significantly influenced by vaccine status of the study patient (Cs-D2, p=0.018), days between vaccination and infection (0.021) and NPS viral load (p=0.011) (Supplementary Table 4). Multivariate Logistic regression analysis adjusted for confounding factors and effect modulation showed Covishield vaccination and duration from the last dose of vaccination to infection were interacting factors, which indicated that a higher percentage of multiple infections in Covishield vaccinated household was likely to be associated with a longer duration from the last dose of vaccine to infection. Individual risk of infection analysis of 95 recruited patients and their 193 household members showed that the fully vaccinated individuals, irrespective of the vaccine type, had a higher risk of infection than unvaccinated (67.4% positivity in fully vaccinated vs 38.1% in unvaccinated patients, p <0.0001, power of 99.46) and remained statistically significant even after adjustment of ages between 18 - 60 years representing the mobile population group (Table 3). All the above analysis indicates a possibility of more infections in the vaccinated than unvaccinated individuals in this cohort.

**Figure 3.**
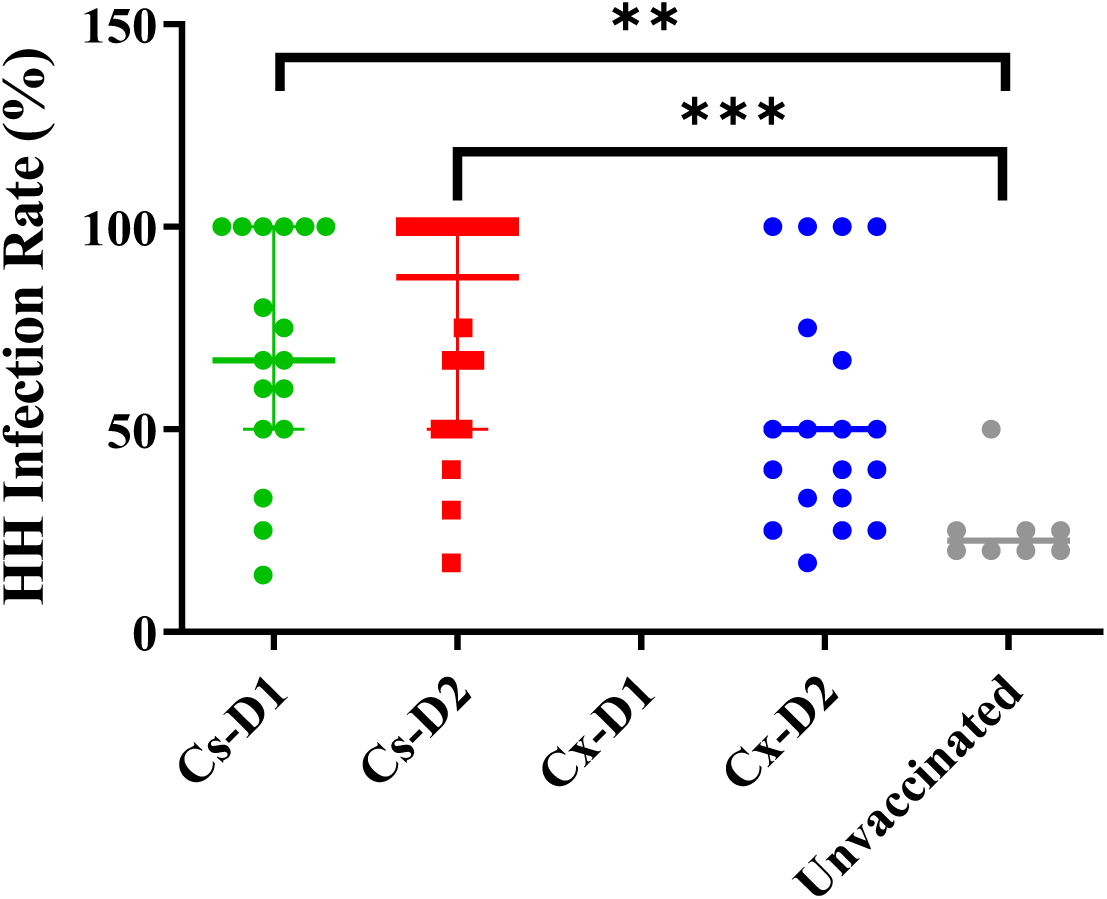
Household infection rate based on the vaccination status of the study patients. The graph excludes household data of multiple study patients with different vaccination status from the same household or if the study patient was living alone. Cs-D1-Covishield Dose 1, Cs-D2-Covishield Dose 2, Cx-D1-COVAXIN Dose 1 and Cx-D2-COVAXIN Dose 2. ** signifies p<0.01 and *** signifies p<0.001.

**Table 3.**
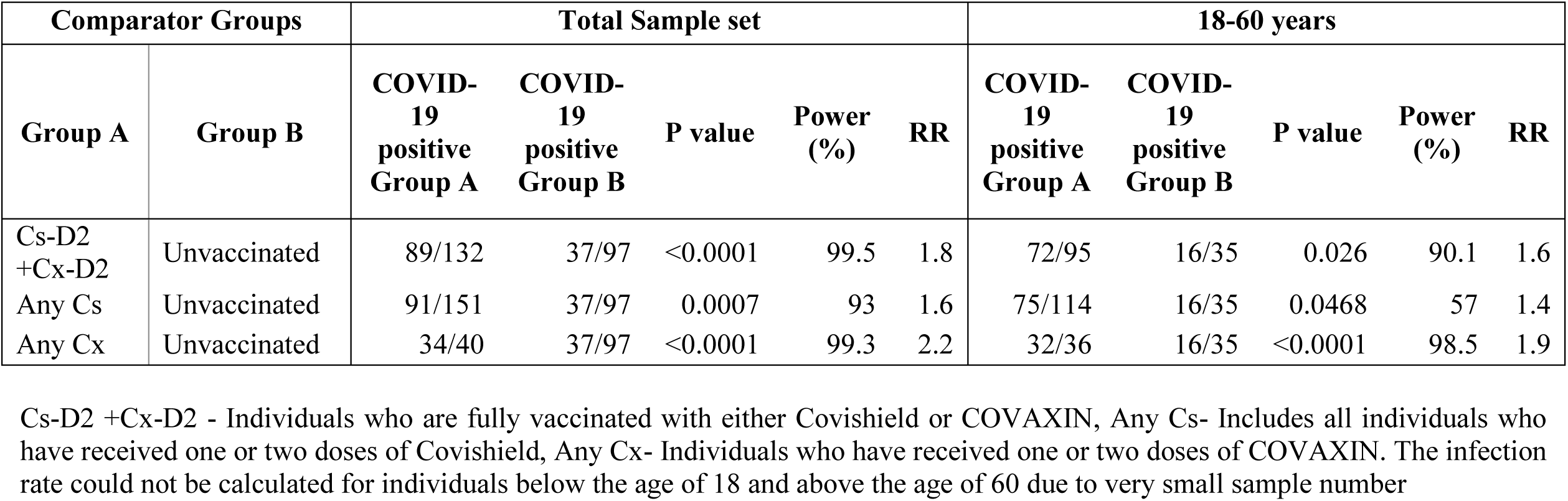
Risk of infection upon exposure to the virus based on vaccination status (n= 288)

| Comparator Groups |  | Total Sample set |  |  |  |  | 18-60 years |  |  |  |  |
| --- | --- | --- | --- | --- | --- | --- | --- | --- | --- | --- | --- |
| Group A | Group B | COVID-19 positive Group A | COVID-19 positive Group B | P value | Power (%) | RR | COVID-19 positive Group A | COVID-19 positive Group B | P value | Power (%) | RR |
| Cs-D2 +Cx-D2 | Unvaccinated | 89/132 | 37/97 | <0.0001 | 99.5 | 1.8 | 72/95 | 16/35 | 0.026 | 90.1 | 1.6 |
| Any Cs | Unvaccinated | 91/151 | 37/97 | 0.0007 | 93 | 1.6 | 75/114 | 16/35 | 0.0468 | 57 | 1.4 |
| Any Cx | Unvaccinated | 34/40 | 37/97 | <0.0001 | 99.3 | 2.2 | 32/36 | 16/35 | <0.0001 | 98.5 | 1.9 |

Cs-D2 +Cx-D2 - Individuals who are fully vaccinated with either Covishield or COVAXIN, Any Cs- Includes all individuals who have received one or two doses of Covishield, Any Cx- Individuals who have received one or two doses of COVAXIN. The infection rate could not be calculated for individuals below the age of 18 and above the age of 60 due to very small sample number

## Discussion

The dynamics of airborne disease transmission is complex, and understanding factors affecting transmission has critical implications for disease control interventions and health policies. A key factor significantly impacting airborne transmission is the viral load in the air, which is directly dependent on the viral load in the respiratory fluids of the infector and the rate at which it is released into the air (17). In this study, we used a simple adapted N95 mask sampling method to measure the impact of Delta variant and COVID-19 vaccination on individuals infected with SARS-CoV-2 to expel the virus into the air and understand its potential contribution to transmission.

### High transmission of the Delta variant is associated with a high viral load in expelled respiratory particles

We showed that 95% of the people infected with the Delta variant expelled virus in their respiratory particles, 2.3-fold more than people infected with the initial SARS-CoV-2 strains in 2020. Further, the magnitude of viral load expelled was significantly more (∼80 fold) with Delta infections. The proportion of high emitters was 3-fold more than in 2020, explaining the high rates of transmission observed with Delta infections. This is congruent with Rowe et al.’s modelling outcome, where they showed that if the Delta strain had a dose of exposure that is 100-fold more than the initial strains, then the probability of infection from it is near 100% (17).

Furthermore, it is well established from epidemiological studies that transmission is heterogeneous and over-dispersed for most infectious diseases, including COVID-19 (18-20). There is a high degree of variability in the ability of infected individuals to cause further transmission, and only a small proportion contributes to 80% of the secondary infections (19). Earlier, we had shown that the mask positivity and proportion of high emitters reflect this variability and the reported secondary attack rates of the initial strains (12), providing a reasonably good indicator for identifying individuals who are likely to cause secondary infections. Even in this study, we observed that 41% of the patients were high emitters, which again correlated to the reported overall secondary attack rate (SAR) of 30.8% (95%CI, 23.5%-39.3%) for Delta infections, derived from multiple household contact studies through a meta-analysis (21). In addition, our results also aligned with the South Korean Delta outbreak study that showed that only 40% of the individuals caused all secondary infections (22). The alignment of the high emitter pattern to epidemiologically observed transmission rates both for initial strain (13) and the Delta strain (current study) further confirms that a high emitter pattern can be used to assess transmission risk. The results highlight that the mask sampling approach used in this study can serve as a good tool to quickly understand transmission risks from new variants like Omicron or any novel respiratory viruses that would be useful in framing policies for patient isolation requirements.

### Transmission risk continues to remain high in a subset of individuals even after eight days of symptom onset

In 2021 until omicron emerged, guidelines across various countries, including India, recommended that mild patients’ isolation be terminated at ten days from symptom onset, provided they did not have fever for about 24 (USA)-72 (India) hours. These guidelines were initially framed based on contact tracing studies and laboratory studies that looked at culturable active viruses from 2020, which showed less than 5% risk for transmission at the late stage (23-25). Even though the Delta variant emerged to be more virulent and transmissible during the second wave in India and subsequently in other countries, the viral shedding dynamics of the infectious virus have just started emerging (9, 26-29). One study that looked at infectious viruses upto15 days showed that infectious viral shedding is longer for the Delta than non-Delta infections (29). In this study, we showed that the probability of patients expelling virus on the tenth day from symptom onset is 65% in individuals who had mild Delta infections (Supplementary Figure 2A); however, the likelihood of being a high emitter was 7% (Supplementary Figure 2C), suggesting that there is a theoretical risk for transmission for Delta even at this late stage of infection. Notably, except for two (0.02%) patients, all patients, including high emitters, had no fever in the follow-up period (Supplementary Table 1). However, the high emitters were significantly associated with having cough as a symptom, suggesting that relying on just fever may overlook the possible risk for transmission. Even though our results show that guidelines of 10 days isolation were applicable in most cases, a subset (7%) may continue to carry high risk and may need longer isolation.

### Vaccinated individuals carry similar transmission risks emphasizing the continued need for masks and social distancing

Another important aspect of the study was the impact of vaccination on potential transmission risk. We noted that irrespective of vaccine taken, the proportion of people who expelled the virus (Table 2) and the magnitude of the viral load (Figure 2) was similar in partial, full and un-vaccinated individuals suggesting that vaccinated individuals are equally likely to expel and transmit the virus. The results of this study are congruent with other studies that reported similar swab viral loads in vaccinated and unvaccinated individuals (7, 9, 30). However, the study differs from other contact tracing and infectious virus studies that showed vaccination reduced transmission risk (8, 9, 27, 28). Our study showed that the households with Covishield vaccinated study patients had a higher proportion of infections in their household (Figure 3). Moreover, at the individual level, the vaccinated persons showed a higher risk for infection acquisition (Table 3). Two reasons may explain these findings-1) we observed that the duration between vaccination and infection influenced the household infection rates, indicating that waning of vaccine-induced immunity with time may cause lower vaccine efficacy against transmission, as noted by other studies (11). The Eyre et al. study (11) noted that Delta transmission among fully vaccinated individuals was similar to unvaccinated by 12 weeks of ChAdOx-nCoV-19 (AZD1222) vaccination. Aligning with this, we noted that the proportion of high emitters was 57 % in patients who got infection 12 weeks after full vaccination vs 35% for <12 weeks. 2) Socio-Behavioural aspects could provide an alternative explanation for the findings. It is pertinent to note that this study was conducted between July and Sep 2021, when the threat of the second wave had reduced, and the vaccination coverage was accelerating in Mumbai among the younger population. The perceived lower risk of acquiring infection upon vaccination, especially among the younger people, may have caused an increased tendency for not following COVID-19 appropriate behaviour by these individuals/households, resulting in more infections than expected. Such behavioral changes have been attributed to the Peltzman effect that suggests people typically adjust their behaviour according to perceived risk (31, 32). Our study highlights the need for appropriate communication of public health messages when interventions are being introduced for better disease control.

## Limitations of the study

An important limitation of the study is that the number of unvaccinated patients recruited was low due to an exponential increase in vaccination rates in the city during the study period, higher institutional care and lesser willingness to give consent for the study among the unvaccinated than vaccinated patients. Moreover, the study did not measure prior infections in the patients through specific antibody testing, which may have influenced patients’ ability to expel the virus and the household infection rates, especially in the unvaccinated group. Lastly, even though the study measures transmission relevant viral load in expelled air, the study estimates viral load from RT-PCR, which does not differentiate between active and inactive virus. Despite these limitations, the study results are consistent with other studies showing that the Delta strain had a propensity for higher transmission, and vaccination may not effectively prevent transmission.

## Conclusion

In conclusion, this study used a simple adapted N95 mask sampling method that captured infectious respiratory particles of COVID-19 patients to demonstrate the impact of the Delta variant and vaccination on potential forward transmission risk. Our study provides biological evidence for increased transmission of the Delta variant and reduced prevention of person-to-person transmission after vaccination. With the constant threat of the emergence of highly transmissible new variants (like Omicron) and the introduction of mass-scale interventions like vaccination, boosters, etc., studies like this become critical to continuously understand transmission patterns of the ongoing pandemic. The tool and methods used in this study have many applications, including-a) understanding the potential transmission risk of any new variants or interventions that can guide the development of patient isolation policies or disease control strategies, b) screening new vaccine or therapeutic candidates for their ability to block transmission.

## Data Availability

All data produced in the present work are contained in the manuscript

## Author Contributions

KS contributed to conceptualization, project development and management, funding acquisition, study design, patient recruitment, data analysis and interpretation, and drafting and revising the manuscript. AS contributed conceptualization, study design, project management, patient recruitment, data analysis and interpretation, drafting and revision of the manuscript. SV, TM, GP and SS contributed to investigations, data acquisition, data management and data visualization. SV also contributed to the revision and editing of the manuscript. VO contributed to conceptualization, providing resources and patient recruitment. PK contributed to sequencing analysis. KN contributed to sequencing resources and revision of the manuscript. DS and MG provided resources and approvals for the successful conduct of the study. NFM contributed to conceptualization, study design, funding acquisition, data interpretation, manuscript revision, and overall supervision.

## Conflict of Intrest Statement

The authors declare no conflict of interest

## Acknowledgements

We are deeply thankful to Mr. Nadir Godrej, Mr. Aditya Berlia, Mr. Rakesh Agarwal, Mr. Pranav Kothari, Mr. Sandeep Chopra, Mr. Anantnarayan Sunderesan and others for their donations. We thank Dr Chandrakant Pawar, Medical Superintendent, Kasturba Hospital for Infectious Diseases, for his initial support towards the study. We deeply appreciate the contribution of Ms. Niharika Shinde, the field researcher who enrolled the patients and collected the samples. Finally, we would like to thank all the participants of this study for their cooperation, patience, and support, without which this study would not have been possible

