## Supplemental Information for "Impact of Delta and Vaccination on SARS-CoV-2 transmission risk: Lessons for Emerging Breakthrough infections"

**Supplementary Table 1 –** Comparison of patient clinical parameters, COVID-19 symptom characteristics stratified based on vaccination status

| Descriptions | Overall | Cs-D1 | Cs-D2 | Cx-D2 | Unvaccinated |
|---|---|---|---|---|---|
| **Clinical parameters** | | | | | |
| Lymphocytes (%) (median + IQR) | 33 (26.7-39.1) | 34 (29.5 - 39.5) | 29 (17-38) | 34.5 (27.5 - 44) | 32 (26-35) |
| Platelets (thou/µL) (median + IQR) | 253 (187-300) | 286 (232 -314) | 240 (177-288) | 236 (174- 307) | 271 (179-509) |
| Hemoglobin (g/dL) (median + IQR) | 13.3 (11.7 -14.2) | 13.5 (12.5 - 14) | 13.4 (11.3 - 15.3) | 13.4 (12.2 -14.1) | 12.5 (11.6-13.6) |
| CRP (mg\L) (median + IQR) | 7.8 (2.9 - 16) | 7.75 (2.8 - 11) | 10.2 (5.3- 20.3) | 6.7 (2.6-18.1) | 8.3 (1.71-13.1) |
| LDH (U/L) (median + IQR) | 246 (173- 295) | 209 (165 - 264) | 256 (168- 280) | 277 (189 -333) | NA |
| **Symptoms at Enrolment** | | | | | |
| Fever | 44 | 16 | 11 | 13 | 4 |
| Cough | 55 | 18 | 13 | 17 | 6 |
| Sore throat | 25 | 10 | 5 | 7 | 1 |
| Loss of Smell/Taste | 51 | 13 | 14 | 17 | 7 |
| Headache | 15 | 4 | 5 | 3 | 3 |
| Body ache/Weakness | 27 | 10 | 4 | 9 | 3 |
| Chills | 7 | 1 | 1 | 4 | 1 |
| Gastric Problems | 5 | 0 | 2 | 1 | 2 |
| Shortness of Breath | 3 | 0 | 0 | 2 | 1 |
| Cold | 25 | 7 | 10 | 6 | 1 |
| **Symptoms at Follow-up** | | | | | |
| Fever | 2 | 0 | 0 | 2 | 0 |
| Cough | 37 | 12 | 7 | 14 | 4 |
| Sore throat | 4 | 0 | 1 | 1 | 2 |
| Loss of Smell/Taste | 37 | 8 | 11 | 15 | 3 |
| Headache | 6 | 1 | 1 | 4 | 0 |
| Body ache/Weakness | 6 | 1 | 0 | 5 | 0 |
| Chills | 1 | 0 | 0 | 1 | 0 |
| Gastric Problems | 6 | 1 | 1 | 4 | 0 |
| Shortness of Breath | 1 | 0 | 0 | 0 | 1 |
| Cold | 12 | 4 | 2 | 5 | 1 |
| **Types of Comorbidities** | | | | | |
| Diabetes | 14 | 0 | 9 | 4 | 1 |
| Hypertension | 9 | 0 | 5 | 2 | 2 |
| Cardiovascular Disease | 9 | 0 | 2 | 6 | 1 |
| Others | 14 | 4 | 2 | 6 | 2 |

All values are represented as the number of patients unless specified otherwise. Cs-D1 – One dose of Covishield, Cs-D2 - Two doses of Covishield and Cx-D2 – Two doses of COVAXIN; Ct value – cycle Threshold value; IQR – interquartile range. Cx-D1 group was not included in the analysis due to low patient number (N=2)

**Supplementary Table 2 – Viral strain of the sequenced patient samples**

| SARS-CoV-2 strain | Number of Patients |
|---|---|
| Delta | 66 |
| Delta Derivative (AY.120) | 9 |

**Supplementary Table 3 – Comparison between Mask High Viral Emitters and Not High Emitters***

| Descriptions | High Emitters (N=37) | Not High Emitters* (N=55) | p-value | Adjusted p-value (Multivariate Analysis) |
|---|---|---|---|---|
| **Age** (median + IQR) | 40 (30-50) | 41 (31-50) | 0.761 | 0.115 |
| **Gender** | | | | |
| Male | 23 | 28 | 0.392 | |
| Female | 14 | 27 | | |
| **Vaccine Status** | | | | |
| Cs-D1 | 7 | 19 | >0.999 | 0.475 |
| Cs-D2 | 14 | 14 | 0.32 | 0.678 |
| Cx-D1 | 1 | 1 | >0.999 | 0.228 |
| Cx-D2 | 11 | 13 | 0.49 | 0.893 |
| Unvaccinated | 4 | 9 | | |
| **Swab Ct Value at Diagnosis** (median + IQR) | 20 (16-24) | 21 (17-25.7) | 0.305 | 0.091 |
| **Swab Viral Load at Recruitment (log Value)** (median + IQR) | 6.5 (5.6 – 7.5) | 4.4 (2.7-5.6) | **<0.0001** | **0.027** Adjusted for Age, Vaccine status, Number of comorbidities, Swab Ct value at Diagnosis, Cough as a symptom, Day's post symptom onset and effect of days post symptom onset and comorbidities on viral load |
| **Number of Symptoms** | 4 (2-5) | 3 (2-4) | 0.341 | |
| **Type of Symptoms** | | | | |
| Fever | 17 | 28 | 0.675 | |
| High | 3 | 4 | >0.999 | |
| Mild | 15 | 25 | 0.673 | |
| Cough | 27 | 28 | **0.038** | 0.334 |
| Sore Throat | 12 | 13 | 0.473 | |
| Loss of Smell and Taste | 16 | 35 | 0.058 | |
| Shortness of Breadth | 2 | 1 | 0.562 | |
| Cold | 12 | 13 | 0.473 | |
| **Number of comorbidities** | 1 (0-1) | 0 | **0.001** | 0.976 |
| **Type of comorbidities** | | | | |
| Diabetes | 9 | 5 | 0.073 | |
| Hypertension | 6 | 4 | 0.193 | |
| CVD | 4 | 1 | 0.153 | |
| Others | 8 | 6 | 0.236 | |
| **Duration between last dose of vaccination to infection** (median + IQR) | 47 (21-87) | 58 (21-77) | 0.707 | |
| **Antivirals taken** | 23 | 32 | 0.829 | |
| **Days post symptom onset** (median + IQR) | 3 (3-4) | 4 (3-5) | **0.023** | 0.566 |
| **Sampling score** (median + IQR) | 7 (6-7.75) | 6 (5-8) | 0.217 | |

*Includes moderate, low, and zero emitters. All values are represented as the number of patients unless specified otherwise. IQR-Interquartile range

**Supplementary Table 4 – Comparison Between Single infection and Multiple infection households**

| Description | Single infection households (N=22) | Multiple infection households (N=67) | p-value | Adjusted p-value (Multivariate Analysis) |
|---|---|---|---|---|
| **Age** (median + IQR) | 39 (30-50) | 40 (31-49) | 0.764 | 0.432 |
| **Gender** | | | | |
| Male | 10 | 39 | 0.331 | |
| Female | 12 | 28 | | |
| **Vaccine status** | | | | |
| Cs-D1 | 7 | 18 | 0.065 | |
| Cs-D2 | 2 | 26 | **0.018** | 0.227 |
| Cx-D2 | 6 | 17 | 0.119 | Adjusted for Age, No of Symptoms, Fever as a symptom, Days between the last dose of vaccination to infection and adjusted for the effect of Days between the last dose of vaccination to infection in CS-D2 patients |
| Unvaccinated | 7 | 4 | | |
| **Days between last dose of vaccination to infection** (median + IQR) | 58 (26-77) | 63 (37-87) | **0.021** | 0.401 |
| **Number of symptoms** | 3 (1-4) | 3 (2-5) | 0.236 | 0.259 |
| **Types of symptoms** | | | | |
| Fever | 9 | 36 | 0.221 | 0.786 |
| Cough | 12 | 17 | 0.67 | |
| Sore throat | 6 | 37 | >0.999 | |
| Gastric problems | 11 | 4 | >0.999 | |
| Cold | 4 | 20 | 0.408 | |
| **Viral load in mask** (Log Value, median + IQR) | 2.2 (1.6-3.7) | 2.8 (1.6-4.1) | 0.371 | |
| **Viral load in Swab** (Log Value, median + IQR**)** | 4.4 (2.-6.1) | 5.6 (4.5-6.6) | **0.011** | |
| **Ct value at diagnosis** (median + IQR) | 21 (17-24) | 20 (16.5-24) | 0.337 | |
| **Family type** | | | | |
| Nuclear | 8 | 34 | 0.326 | |
| Joint | 14 | 33 | | |
| **Days post symptom onset** (median + IQR) | 4 (3-5) | 4 (3-5) | 0.495 | |
| **Antivirals Taken** | 12 | 43 | 0.455 | |

Single infection Household – Only study patient infected with SARS-CoV-2, multiple infection Household – Study patient + Household member/s infected with SARS-CoV-2, IQR-Interquartile range

**Supplementary Figure 1**

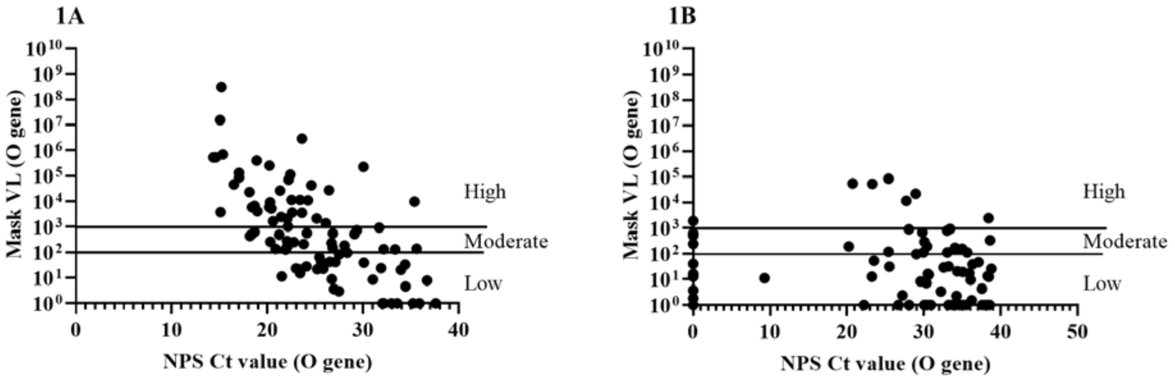


**Supplementary Figure 1:** Scatter plot depicting the NPS Ct value on X-axis and mask viral load on Y-axis **1A-** at Enrolment and **1B** – Follow up

**Supplementary Figure 2**

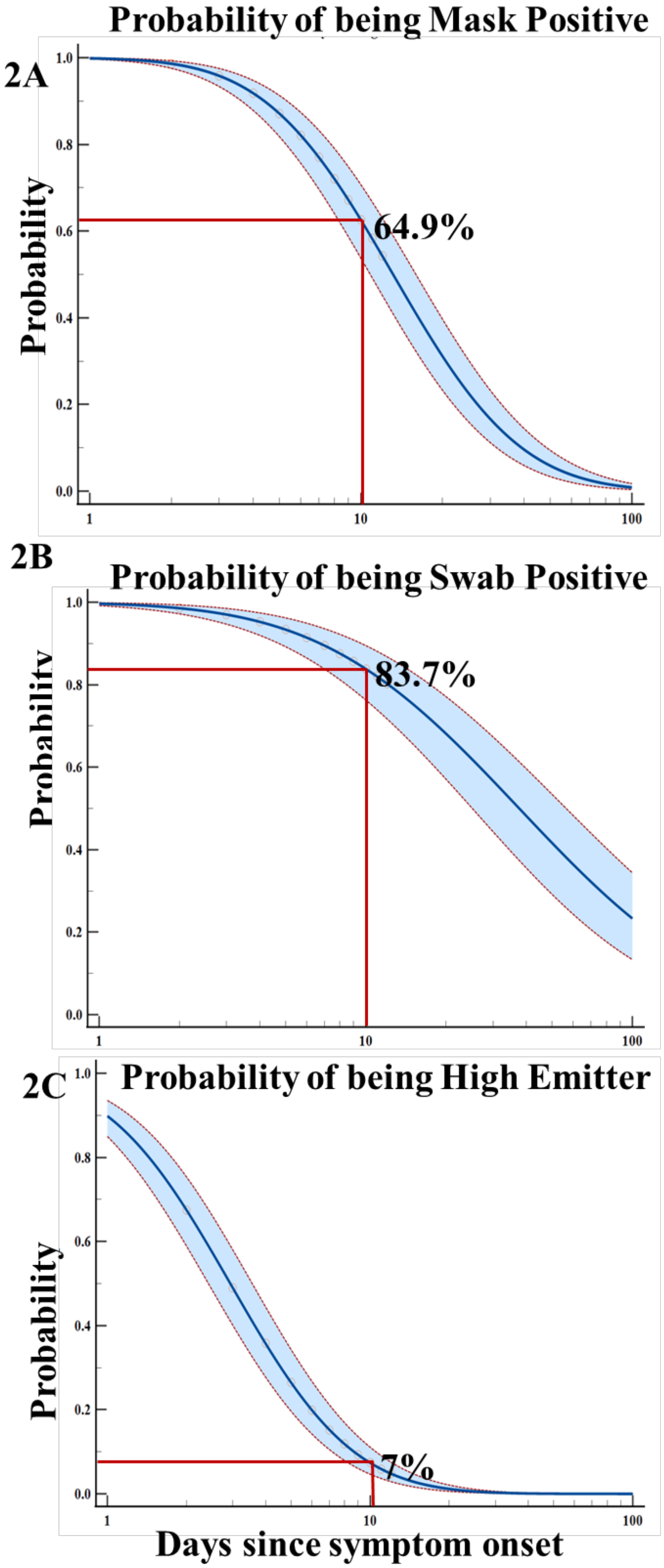


**Supplementary Figure 2 – Probit Analysis** – The graphs describe the probability of being mask positive (2A), swab positive (2B) or a high emitter (2C) on any day after the symptom onset of SARS-CoV-2 infection as calculated by Probit (MedCalc software, 20.019). The red lines highlight the probability of being mask (2A), swab (2B) positive or high emitter (2C) tenth day from symptom onset.
